# Interaction-based Mendelian randomization with measured and unmeasured gene-by-covariate interactions

**DOI:** 10.1101/2020.07.27.20162909

**Authors:** Wes Spiller, Fernando Pires Hartwig, Eleanor Sanderson, George Davey Smith, Jack Bowden

## Abstract

Studies leveraging gene-environment (GxE) interactions within Mendelian randomization (MR) analyses have prompted the emergence of two methodologies: MR-GxE and MR-GENIUS. Such methods are attractive in allowing for pleiotropic bias to be corrected when using individual instruments. Specifically, MR-GxE requires an interaction to be explicitly identified, while MR-GENIUS does not. We critically examine the assumptions of MR-GxE and MR-GENIUS, and propose sensitivity analyses to evaluate their performance. Finally, we explore the association between body mass index (BMI) and systolic blood pressure (SBP) using data from the UK Biobank. We find both approaches share similar assumptions, though differences between the approaches lend themselves to differing research settings. Where interactions are identified, MR-GxE relies on weaker assumptions and allows for further sensitivity analyses. MR-GENIUS circumvents the need to identify interactions, but relies on the MR-GxE assumptions holding globally. Through applied analyses we find evidence of a positive effect of BMI upon SBP.

---

Mendelian randomization (MR) is an epidemiological approach applied to observational data, in which genetic variants are used as instrumental variables (IVs) in order to estimate the causal effect of a modifiable exposure on a downstream outcome^1^. It encompasses a wide range of statistical methods, and typically relies upon three key assumptions to test for causality. A suitable genetic IV is strongly associated with the exposure of interest (IV assumption 1), independent of all confounders of the exposure and outcome, as well as confounders of the IV and outcome (IV2), and independent of the outcome when conditioning on the exposure and all confounders of the exposure and outcome (IV3)^1, 2^. Direct associations between a genetic instrument and the outcome of interest are defined as *horizontal pleiotropic* pathways^3^. Associations which violate IV2-3 represent the set of associations between the genetic variant and the outcome which are unrelated to the exposure, introducing bias into estimates of causal effect^4^. Pleiotropic associations are also believed to be potentially more likely when examining complex phenotypes in MR analyses, inducing bias in causal effect estimates. As a consequence, pleiotropy robust methods have been a central research focus in MR methods development^2, 5, 6^.

One strategy to mitigate the problem of pleiotropic bias is to leverage variation in instrument strength across one or more covariates within a target population, represented as a gene-by-covariate interaction^7-9^. Intuitively, if it were possible to identify a subgroup of the population wherein the instrument and exposure are independent (i.e. a ‘no-relevance group’), it follows that, in the absence of pleiotropy, the corresponding instrument and outcome should also be independent. A non-zero instrument-outcome association in such a situation therefore indicates that pleiotropy is present^7, 10, 11^. Empirically, however, no-relevance groups of sufficient size are rarely observed, prompting the development of more sophisticated approaches that leverage statistical assumptions to extrapolate back to a hypothetical no-relevance group. Two such methods are MR using Gene-by-Environment interactions (MR-GxE) and MR G-Estimation under No Interaction with Unmeasured Selection (MR-GENIUS)^7, 12^. MR-GxE uses an explicit gene-by-covariate interaction to estimate causal effects. MR-GENIUS does not require observed interacting covariates, but implicitly leverages all possible gene-by-covariate interactions that induce a dependence between the instrument and the exposure variance. There is, however, little guidance as to the relative strengths and limitations of each approach.

In this paper we implement MR-GxE in an individual level data setting, and critically evaluate the performance of MR-GxE and MR-GENIUS through simulation. We demonstrate how both approaches share similar underlying assumptions and are best applied in differing research settings, specifically whether a suitably ‘strong’ gene-by-environment interaction is, or is not, directly observed. When such a covariate is observed MR-GxE allows for a number sensitivity analyses to be exploited, with related assumptions applying only to the adopted interaction. In contrast, MR-GENIUS requires assumptions to hold across the entire set of potential interactions. Finally, we perform an applied analysis estimating the effect of adiposity upon systolic blood pressure (SBP) using data from the UK Biobank, finding evidence of a positive association between adiposity and SBP using both MR-GxE and MR-GENIUS.

## Results

### Overview of MR-GxE and MR-GENIUS

The MR-GxE and MR-GENIUS approaches use differences in instrument strength across one or more covariates to estimate and correct for pleiotropic bias^7^. For *i* ∈ {1,2, …, *N*} observations, let *G*_*i*_ denote a single genetic instrument for an exposure *X*_*i*_, and let *Y*_*i*_ represent the outcome of interest. Further, assume there exists an unmeasured confounder *U*_*i*_ of *X*_*i*_ and *Y*_*i*_, and a set of interaction covariates ***Z***_*i*_ = {*Z*_1_ … *Z*_*K*_} across which the instrument-exposure association varies. In order to make our ideas concrete, we now define an underlying data generating model for a continuous exposure and outcome, which are themselves a function of genetic variants *G*_*i*_, *Z*_*i*_ and *U*_*i*_.

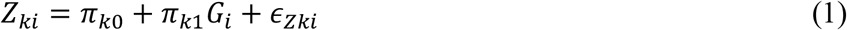

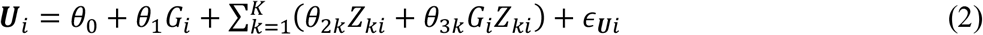

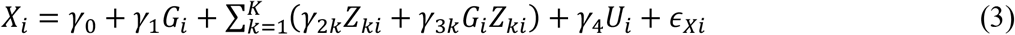

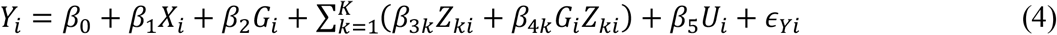

In equations (1-4), the *ϵ*_(.*i*)_ terms represent independent error terms, and relationships with reference to a single interaction covariate *Z*_*ki*_ are illustrated in Figure 1 wherein *G*_*i*_, *Z*_*ki*_, and *U*_*i*_ are assumed independent for clarity.

**Figure 1:**
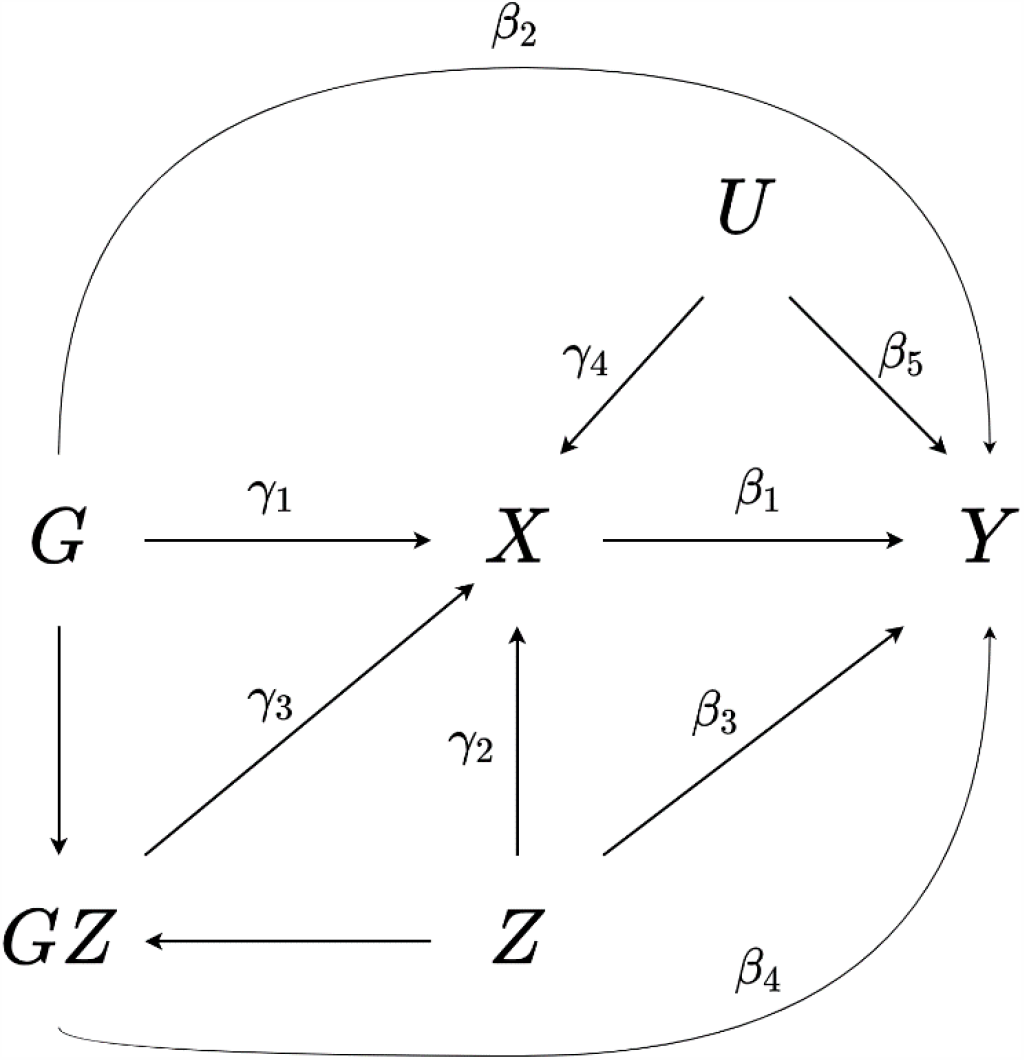
A directed acyclic graph showing the relationship between a genetic instrument *G*, interaction covariate *Z*, exposure *X*, outcome *Y*, and one or more confounders *U. GZ* denotes the interaction *G* × *Z*, and *G, Z*, and *U* are assumed independent for clarity.

The MR-GxE approach requires an interaction covariate (*Z*_*i*_) to be explicitly observed, in contrast to MR-GENIUS which leverages variance differences for a given exposure (*X*_*i*_) across subgroups of a genetic instrument (*G*_*i*_). Such variances differences implicitly rely on the presence of one or more unmeasured interaction covariates. For both approaches, unbiased estimation of causal effects requires three assumptions to be satisfied, summarised as assumptions GxE1-3 below. A suitable interaction (*G*_*i*_*Z*_*i*_) is:

GxE1: Strongly associated with the exposure of interest.

GxE2: Independent of confounders of the exposure and outcome.

GxE3: Not directly associated with the outcome of interest.

For MR-GxE each assumption is framed with reference to a single observed interaction covariate. In contrast, MR-GENIUS requires assumptions GxE1-3 to hold globally across the set of observed and unobserved gene-by-covariate interactions in the sample (*G*_*i*_***Z***_*i*_). For example, GxE1 is satisfied with respect to MR-GENIUS when the average interaction strength for all possible interactions (*γ*_3*K*_) is non-zero across the sample.

MR-GxE is implemented by using a gene-by-covariate interaction as an instrument within a two-stage least squares (TSLS) framework. In the first-stage model (equation 5), the exposure is regressed upon a genetic instrument and observed interaction covariate including an interaction term. The second-stage model (equation 6) then regresses the outcome upon the genetic instrument, interaction covariate, and fitted values for the exposure obtained using the first-stage model.

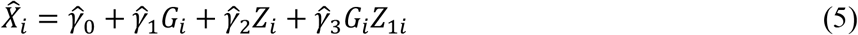

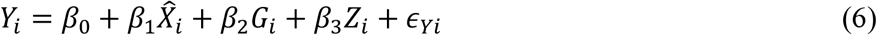

This returns a causal effect estimate 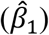, as well as an estimate of pleiotropic effect as the coefficient of the genetic instrument 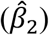 in the second-stage model (see Methods). MR-GENIUS is implemented by first performing a simple regression of the exposure upon the genetic instrument and obtaining a set of residuals 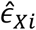. These residuals are then used in conjunction with the genetic instrument and incorporated within a TSLS model as a single instrument for the exposure (see Methods)^12^.

### MR-GxE sensitivity analyses and simulations

By explicitly defining an interaction-covariate, it is possible to perform a range of sensitivity analyses which evaluate assumptions GxE1-3. MR-GxE is reliant upon a strong first-stage interaction (GxE1), and where this is not the case estimates will exhibit *weak instrument bias* (see Methods). The first-stage F-statistic for the interaction term in the first-stage model can be used to identify candidate gene-by-covariate interactions, in a similar fashion to utilising GWASs to identify genetic variants associated with a phenotype of interest. Identifying candidate interactions is performed by fitting the first-stage MR-GxE model for each candidate interaction covariate *Z*_*ki*_ and calculating the F-statistic with respect to *G*_*i*_*Z*_*ki*_ (see equation 5 and Methods)^9^. Applying a Bonferroni multiple testing correction, it is then possible to plot and identify interaction covariates for which the MR-GxE model is identified^13^.

To illustrate the role of interaction strength in MR-GxE and MR-GENIUS analyses, we present two simulated examples using the data generating models given in equations 1-5. Scenario 1 uses a single simulated dataset to demonstrate how sufficiently strong interactions can in principle be identified and visualised by evaluating the F-statistic for the interaction term in the first-stage model (equation 5). We use a sample size of 10,000 including *K* = 100 potential interaction covariates. Of the 100 interaction covariates, 10 were designated to have a first-stage interaction, assigning a value for *γ*_3*k*_ sampled from a normal distribution with mean 2 and standard deviation 0.5. Remaining interaction covariates were assigned values for *γ*_3*k*_ sampled from a normal distribution with mean 0 and standard deviation 0.01. The complete set of interaction covariates *Z*_*Ki*_ were generated so as to be independent of *G*_*i*_, such that *π*_*k*1_ = 0 in equation (1). Scenario 2 illustrates how weak instrument bias results in biased estimates of causal effect. This is achieved using the same data generating model as in simulation 1, varying the strength of the first-stage interaction to demonstrate the effect of weak instrument bias. Causal effect estimates represent the mean estimate for a given mean F-statistic across a total of 5000 iterations, and were obtained using a separate sample of 10,000 observations.

The results of simulations 1 and 2 are presented in Table 1 and Figure 2 below. For simulation 1, Figure 2a shows how a scatter plot can be constructed in a similar fashion to a Manhattan plot in GWAS analyses. In this case, a Bonferroni multiple testing correction can be visualised using a solid horizontal line. From Table 1 it can be seen that using individual interaction covariates within the MR-GxE framework provides results comparable to MR-GENIUS when the assumptions of both approaches are satisfied. Simulation 2 demonstrates how causal effect estimates exhibit both bias and a loss of precision as interaction strength decreases, as shown using a forest plot in Figure 2b and presented in Table 1.

**Table 1:** Simulated results and effect estimates for subset of interaction (denoted *Z*) identified from Figure 2a (simulation 1), and results illustrating direction of bias under weak instrument (interaction) (simulation 2).

| Covariate | Simulation 1 |  | Simulation 2 |  |
| --- | --- | --- | --- | --- |
| | $\hat{\beta}_1$ (CI) | F-statistic (p-value) | Mean F-statistic | $\bar{\beta}_1$ (CI) |
| $Z_5$ | 1.003 (0.99-1.01) | 341.7 (<0.001) | 8.7 | 0.86 (-0.91, 2.63) |
| $Z_{21}$ | 1.004 (0.99-1.02) | 111.5 (<0.001) | 13.3 | 0.93 (0.22, 1.65) |
| $Z_{30}$ | 1.008 (1.00-1.02) | 443.8 (<0.001) | 19.1 | 0.97 (0.44, 1.50) |
| $Z_{36}$ | 1.007 (1.00-1.02) | 264.4 (<0.001) | 25.5 | 0.97 (0.54, 1.41) |
| $Z_{37}$ | 1.006 (1.00-1.01) | 406.5 (<0.001) | 32.9 | 0.98 (0.61, 1.35) |
| $Z_{54}$ | 0.997 (0.99-1.01) | 222.8 (<0.001) | 41.7 | 0.99 (0.66, 1.31) |
| $Z_{64}$ | 1.002 (0.99-1.01) | 329.7 (<0.001) | 51.0 | 0.99 (0.70, 1.28) |
| $Z_{77}$ | 1.001 (0.98-1.02) | 76.5 (<0.001) | 60.9 | 0.99 (0.73, 1.25) |
| $Z_{78}$ | 1.001 (0.99-1.01) | 250.9 (<0.001) | 72.8 | 0.99 (0.75, 1.23) |
| $Z_{96}$ | 1.015 (1.01-1.02) | 376.2 (<0.001) | 85.4 | 0.99 (0.77, 1.21) |
| **MR-GENIUS** | **1.008 (1.00-1.02)** | **280.0 (<0.001) <sup>a</sup>** | - | - |

<sup>a</sup> Identification test using Breusch-Pagan test for heteroskedasticity.

**Figure 2:**
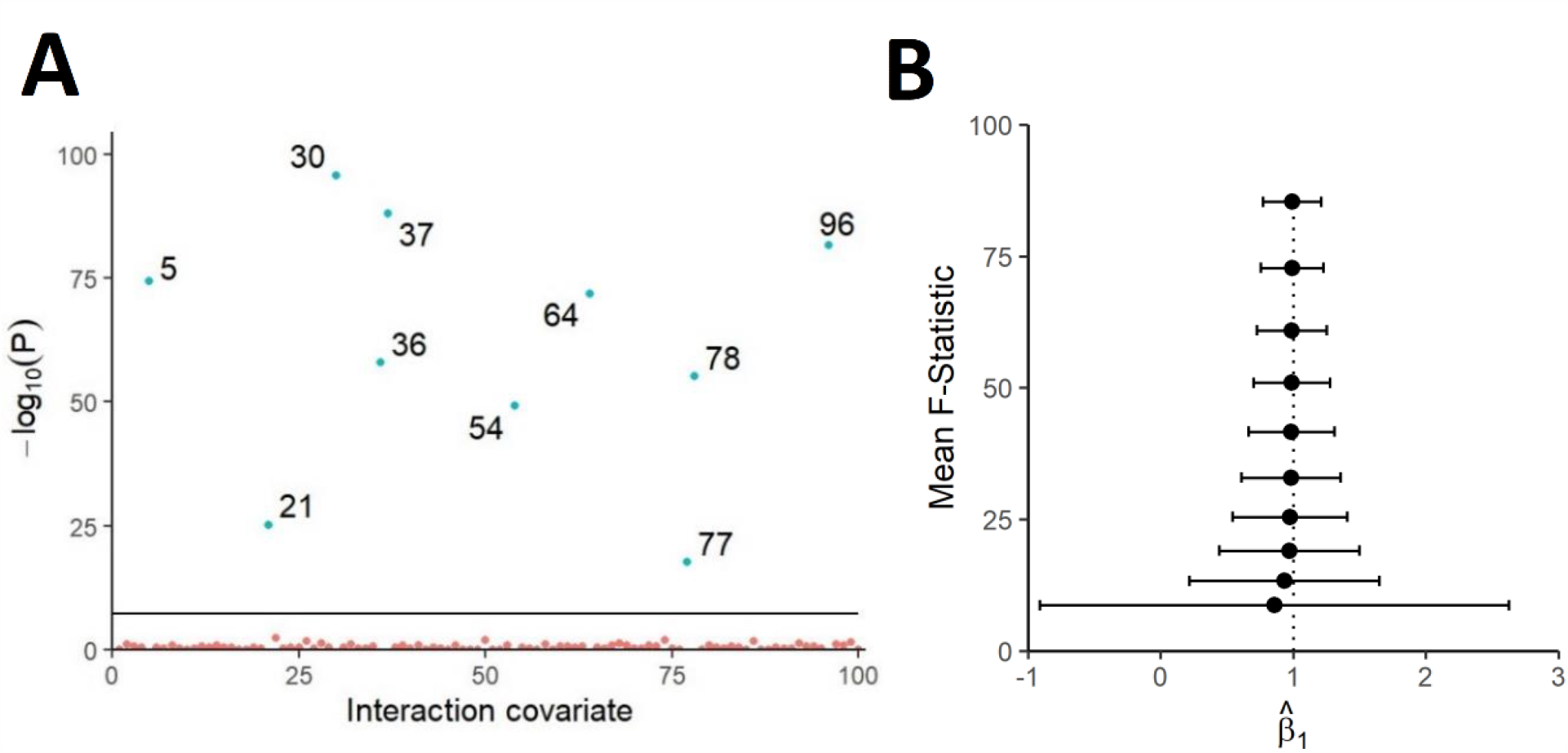
Panel A shows a scatter plot of − log_10_(*p* − *value*) for the first-stage F-statistic across the set of 100 potential interaction covariates for a single simulated data set. Panel B shows a forest plot of mean causal effect estimates and confidence intervals under varying mean interaction strength. A solid horizontal line is included representing the Bonferroni correction threshold for statistical significance in panel A, while the dotted vertical line in panel B represents the true causal effect *β*_1_ = 1.

MR-GENIUS requires the set of unobserved interactions to be sufficiently strong globally to provide sufficiently precise estimates of causal effect. This can be shown in a further simulated example under scenario 1 above, with the addition that the proportion of non-zero interactions is varied within the sample. Table 2 contrasts estimates obtained using a *single* valid and explicitly defined interaction using MR-GxE with MR-GENIUS estimates implicitly using the *entire* interaction set. It can be seen that as the mean interaction strength increases across the set of *K* covariates the precision of the MR-GENIUS approach improves. This highlights the utility of the MR-GxE approach where interaction covariates are readily identifiable, and the flexibility of the MR-GENIUS approach in the absence of observed gene-by-covariate interactions.

**Table 2:** Simulated results using differing proportions of non-zero interaction covariates.

| Proportion of $\gamma_{3K} \neq 0$ | Mean $\gamma_{3K}$ | MR-GxE $\hat{\beta}_1$<br>(CI) | MR-GENIUS $\hat{\beta}_1$<br>(CI) | BP-Test p-value |
| --- | --- | --- | --- | --- |
| 1% | 0.020 | 1.000 (0.99, 1.01) | 1.006 (0.87, 1.14) | 0.083 |
| 5% | 0.100 | 1.000 (0.99, 1.01) | 1.001 (0.94, 1.06) | <0.001 |
| 10% | 0.200 | 1.000 (0.99, 1.01) | 1.002 (0.96, 1.05) | <0.001 |
| 25% | 0.500 | 1.000 (0.99, 1.01) | 1.001 (0.96, 1.04) | <0.001 |
| 50% | 1.000 | 1.000 (0.99, 1.01) | 1.001 (0.96, 1.04) | 0.001 |
| 75% | 1.499 | 1.000 (0.99, 1.01) | 1.001 (0.96, 1.04) | 0.004 |
| 100% | 2.004 | 1.000 (0.99, 1.01) | 1.001 (0.96, 1.04) | 0.006 |

Assumption GxE2 requires the gene-by-covariate interaction to be independent of common causes and confounders of the exposure and outcome. This is equivalent to instrument exogeneity in the conventional MR setting, though it should be noted that associations between the instrument *G*_*i*_ and the interaction covariate *Z*_*i*_ do not *necessarily* introduce bias into MR-GxE estimates. This is most likely the result of problematic confounding structures between *G*_*i*_, *Z*_*i*_, and *U*_*i*_, and consequently it is possible to estimate the correlation between *G*_*i*_ and *Z*_*i*_, with independence serving as evidence of GxE2 being satisfied. However, it should be highlighted such independence does not necessary imply that GxE2 is satisfied, but an observed correlation can highlight potential issues that warrant further consideration (see Methods).

The third assumption GxE3 requires pleiotropic effects of *G*_*i*_ upon *Y*_*i*_ to remain constant across values of *Z*_*i*_, with the gene-by-covariate interaction being independent of *Y*_*i*_ when conditioning on *X*_*i*_. Where this is not the case estimates of causal effect will exhibit bias in the direction of *β*_4_ in a similar fashion to horizontal pleiotropic bias in univariate MR analyses. By reframing MR-GxE within a TSLS framework, it is possible to apply tests of over-identification to evaluate the constant pleiotropy assumption, though this is not possible where only one instrument is available, for example, a single genetic variant. In cases where the single instrument is comprised of many instruments, such as a polygenic risk score, it is possible to examine different configurations of instruments iteratively using MR-GxE and assess heterogeneity in the set of MR-GxE estimates obtained from each iteration. These subsets of instruments are hereafter referred to as *sub-instruments*.

In this scenario, a Sargan test can be used to compare different MR-GxE estimates of the same causal parameter (the coefficient of *X*_*i*_ in equation 6 – i.e., *β*_1_), assuming we have more instruments than we need to consistently estimate the parameter^14^. However, it is important to note that in applying this test it is crucial for each of the sub-instruments to be sufficiently strong to overcome weak instrument bias, though practically the test can be applied where weak interactions are present if assessing the strength of individual instruments of interest.

To demonstrate the impact of GxE3 violation, as well as the utility of employing an adapted Sargan test as a sensitivity analysis, we present a further simulation shown in Table 3. In this case, a score analogous to a PRS was used as a single IV, comprised of 10 individual sub-instruments of approximately equal strength. Mirroring the previous simulated example, the true causal effect was defined as *β*_1_ = 1 with a horizontal pleiotropic effect *β*_2_ = 0.05. Sub-instruments violating assumption GxE3 were estimated to have a value *β*_4_ = 0.2, varying the proportion of invalid sub-instruments across the set of simulated models. The mean F-statistic for the simulations is 698.5 (Breusch-Pagan 253.97 *p* < 0.001), and MR-GENIUS estimates are presented for comparison.

**Table 3:** Simulated results illustrating GxE3 and Sargan test.

| Proportion of valid interactions | Mean $\beta_4$ | MR-GxE $\hat{\beta}_1$ (CI) | GENIUS $\hat{\beta}_1$ (CI) | Sargan p-value |
| --- | --- | --- | --- | --- |
| **100%** | 0 | 0.974 (0.92, 1.08) | 1.000 (0.92, 1.08) | 0.501 |
| **80%** | 0.04 | 1.384 (1.29, 1.44) | 1.179 (1.10, 1.26) | <0.001 |
| **60%** | 0.08 | 1.736 (1.66, 1.80) | 1.367 (1.26, 1.47) | <0.001 |
| **40%** | 0.12 | 2.068 (2.01, 2.18) | 1.544 (1.40, 1.69) | <0.001 |
| **20%** | 0.16 | 2.470 (2.35, 2.56) | 1.727 (1.543, 1.910) | <0.001 |
| **0%** | 0.2 | 2.742 (2.69, 2.94) | 1.908 (1.68, 2.14) | 0.502 |

As shown in Table 3, both MR-GxE and MR-GENIUS produce biased causal effect estimates when the constant pleiotropy assumption is violated. Violation of the constant pleiotropy assumption is also detected by applying a Sargan test, provided all sub-instruments do not identically violate GxE3 such that *β*_4*k*_ is constant. As the Sargan test relies upon at least one instrument being valid, identical violation of assumption GxE3 would also violate the assumptions of the conventional Sargan approach.

### Estimating the effect of adiposity on systolic blood pressure within the UK Biobank

To demonstrate each of the sensitivity analyses previously described, we performed MR analyses estimating the causal effect of adiposity (measured using BMI) on SBP using data from the UK Biobank. This serves as a re-examination of the original applied example in Spiller et al (2019) who first proposed the MR GxE model^7^. Here we go further by evaluating each underlying assumption using the diagnostic tools described above, and contrasting the results with MR-GENIUS^7, 12^. After performing quality control, removing participants with missing data, and restricting the sample to unrelated individuals of European ancestry, a total of 358,928 participants were included in the analyses.

MR-GxE was implemented by constructing a weighted PRS informed using genetic variants previously identified from the GIANT consortium^15^. As the GIANT consortium represents a subset of the most recent UK Biobank release, subsequent analyses have been conducted in a one-sample framework. A total of 95 independent genetic variants were used after performing linkage disequilibrium (LD) pruning, and removing tri-allelic or palindromic variants. Finally, we standardized BMI, SBP, and the weighted PRS using a z-score transformation prior to performing analyses. In previous work we found evidence of a positive association between BMI and SBP using OLS and TSLS regression approaches^7, 16-18^.

Initially, a discovery subset (N=100,000) was randomly sampled from the UK Biobank data for use in identifying interactions for MRGxE analyses. Causal effect estimates and sensitivity analyses were performed using the remaining data. Candidate gene-by-covariate interactions were detected by estimating the first-stage F-statistic for 576 candidate interaction covariates within the UK Biobank. After applying a multiple testing correction, the 20 interaction covariates with the strongest association were selected and utilised in subsequent analyses. Table 4 shows MR-GxE estimates of causal effect and corresponding sensitivity analyses with respect to each interaction covariate. The strength of each interaction across the set of candidate interaction covariates is illustrated in Figure 3, where annotations give the UK Biobank field ID for each interaction covariate.

**Table 4:** MR-GxE estimates and sensitivity analyses using each candidate interaction covariate and MR-GENIUS. The F-statistic reported refers to the F-statistic for the interaction term using the PRS, while 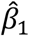 corresponds to the causal effect estimates obtained using each exposure.

| Covariate<br>(UK Biobank Field ID) | F-statistic | $\hat{\beta}_1$<br>(p-value) | $\rho(G, Z)^a$<br>(p-value) | Sargan<sup>b</sup><br>(p-value) | Mean<br>F<sup>c</sup> |
| --- | --- | --- | --- | --- | --- |
| Waist circumference<br>(f.48.0.0) | 182.86 | -0.524<br>($< 0.001$) | 0.103<br>($< 0.001$) | 7.531<br>(0.481) | 79.40 |
| Weight (kg)<br>(f.21002.0.0) | 123.16 | -0.687<br>($< 0.001$) | 0.119<br>($< 0.001$) | 9.342<br>(0.314) | 48.79 |
| Diabetes diagnosis<br>(f.2443.0.0) | 54.22 | -0.065<br>(0.470) | 0.020<br>($< 0.001$) | 12.19<br>(0.143) | 41.22 |
| Alcohol intake frequency<br>(f.1558.0.0) | 50.65 | 0.163<br>(0.006) | 0.001<br>(0.526) | 5.69<br>(0.682) | 41.62 |
| Physical activity (vigorous)<br>(f.904.0.0) | 42.10 | 0.017<br>(0.862) | 0.004<br>(0.003) | 20.40<br>(0.009) | 17.03 |
| Vascular/ heart problem diagnosis<br>(f.6150.0.0) | 33.65 | -0.446<br>($< 0.001$) | 0.028<br>($< 0.001$) | 7.22<br>(0.513) | 16.76 |
| Time number displayed during memory test<br>(f.4253.0.5) | 28.42 | -2.155<br>(0.333) | 0.015<br>(0.002) | 14.54<br>(0.069) | 13.51 |
| Number of days per week walked 10+ mins<br>(f.864.0.0) | 27.87 | 0.208<br>(0.011) | 0.001<br>(0.705) | 6.87<br>(0.551) | 18.98 |
| DBP (automated, baseline)<br>(f.4079.0.0) | 26.45 | -0.324<br>($< 0.001$) | 0.020<br>($< 0.001$) | 6.39<br>(0.603) | 16.32 |
| Physical activity (moderate)<br>(f.884.0.0) | 23.60 | 0.165<br>(0.107) | 0.001<br>(0.324) | 3.66<br>(0.886) | 14.27 |
| Townsend deprivation index<br>(f.189.0.0) | 23.01 | 0.108<br>(0.489) | -0.016<br>($< 0.001$) | 9.24<br>(0.323) | 16.80 |
| Comparative body size at age 10<br>(f.1687.0.0) | 20.65 | 0.283<br>(0.004) | 0.048<br>($< 0.001$) | 9.94<br>(0.269) | 14.62 |
| Time to complete pair matching activity<br>(f.400.0.2) | 20.49 | 0.052<br>(0.689) | -0.007<br>($< 0.001$) | 29.50<br>($< 0.001$) | 11.84 |
| Pulse rate<br>(f.4194.0.0) | 20.45 | 0.031<br>(0.873) | -0.010<br>($< 0.001$) | 13.78<br>(0.088) | 4.77 |
| Time watching television<br>(f.1070.0.0) | 20.01 | -0.140<br>(0.211) | 0.017<br>($< 0.001$) | 14.83<br>(0.063) | 15.08 |
| DBP (automated, follow-up)<br>(f.4079.0.1) | 19.55 | -0.501<br>($< 0.001$) | 0.016<br>($< 0.001$) | 6.37<br>(0.606) | 11.72 |
| Own or rent accommodation<br>(f.680.0.0) | 18.41 | 0.078<br>(0.607) | -0.006<br>($< 0.001$) | 19.84<br>(0.011) | 10.13 |
| Age at assessment<br>(f.21003.0.0) | 18.15 | 0.697<br>($< 0.001$) | 0.013<br>($< 0.001$) | 17.28<br>(0.027) | 14.03 |
| Birthweight known<br>(f.120.0.0) | 17.93 | 0.067<br>(0.851) | -0.014<br>($< 0.001$) | 14.16<br>(0.078) | 4.94 |
| Year of birth<br>(f.34.0.0) | 15.85 | 0.710<br>($< 0.001$) | -0.014<br>($< 0.001$) | 17.12<br>(0.029) | 13.94 |
| **Observational** | - | **0.1864**<br>($< 0.001$) | - | - | - |
| **Two-stage least squares** | **7776.52** | **0.1303**<br>($< 0.001$) | - | - | - |
| **MR-GENIUS** | **1332.7**<br>($< 0.001$)<sup>d</sup> | **0.034**<br>(0.009) | - | - | - |

Bonferroni testing correction p-value $< 5 \times 10^{-5}$ .
<sup>a</sup> $\rho(G, Z)$ represents the correlation between the PRS and interaction covariate.
<sup>b</sup> Sargan shows the results to overidentification tests using multiple sub-instruments.
<sup>c</sup> The mean F-statistic for the sub-instruments is given under Mean F.
<sup>d</sup> BP Heterogeneity Test

**Figure 3:**
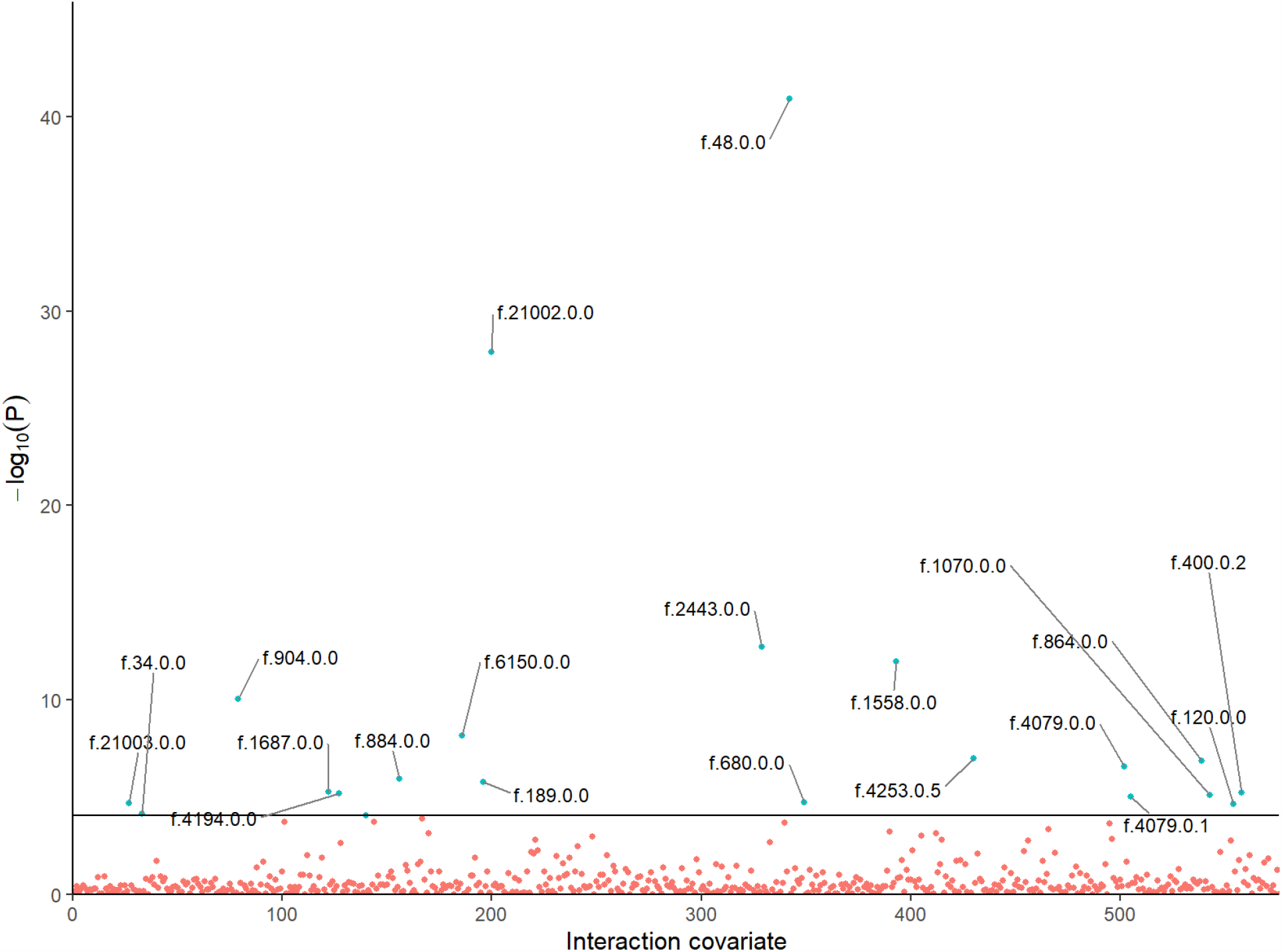
A scatter plot showing the first-stage F-statistics for instrument-by-covariate interactions using data from UK Biobank. A horizontal line is included representing the Bonferroni correction for statistical significance. For clarity, blue points represent interactions identified after multiple testing. The 20 strongest interactions have been annotated using their UK Biobank field identification number.

To assess assumption GxE2, we created 9 sub-instruments sampling from the 95 SNPs used to create the initial PRS instrument. Fitting the MR-GxE model using multiple sub-instruments allows for overidentification tests to be performed, testing the extent to which causal effect estimates differ when using individual sub-instruments. In each case, a failure to reject the null can be considered to be evidence of interaction exogeneity as previously outlined. To implement this approach, the set of SNPs were randomly assorted into 9 sub-instruments of approximately equal strength, quantified using the F-statistic with respect to BMI. Repeating this procedure using sub-instruments containing differing SNPs yielded similar results. We also present the mean F-statistic across the set of sub-instruments to emphasise their strength.

As shown in Table 4, there exists substantial disagreement across the range of selected interaction covariates, suggesting that one or more violate underlying assumptions of the MR-GxE approach.

Considering assumption GxE2, several of the identified gene-by-covariate interactions are proxy measures of adiposity, specifically waist circumference, weight in kilograms, and comparative body size at age 10. Such interaction covariates are often problematic, as associations between the genetic variants and the interaction can result in collider bias where the interaction covariate is downstream of the exposure (see Methods). In this case, higher estimates of *ρ* (*G, Z*) for these variables supports this interpretation, and their subsequent exclusion from further analyses. A similar argument can also be made with respect to interaction covariates downstream of BMI, including diabetes diagnosis, vascular/heart problem diagnosis, and diastolic blood pressure (DBP).

By applying Sargan tests, a number of interaction covariates related to cognition, physical activity, and age appear to violate assumption GxE3. This could be explained by the gene-by-covariate interactions relating to one or more underlying risk factors, which are not adjusted for in the corresponding MR-GxE models.

After applying sensitivity analyses, three interaction covariates can be identified as appropriate choices for estimation using MR-GxE. This selection was made using Sargan test and correlation p-value thresholds of < 0.0025, applying a multiple testing correction. Selected covariates include alcohol intake frequency and physical activity, both days walked and moderate levels of exercise. Considering alcohol intake and physical activity, the lack of a substantial correlation between each interaction covariate and the PRS suggests that violation of GxE2 is unlikely.

In previous work Townsend deprivation index (TDI) was selected as an interaction covariate in a summary MR-GxE analysis and returned estimates in agreement with both alcohol consumption and physical activity measures identified above. However, it is important to note that TDI shows evidence of a non-zero instrument-interaction covariate correlation, potentially highlighting a violation of assumption GxE2. This can be explained by TDI being plausibly downstream of both BMI and the instrument, representing situation in which the correlation does not invalidate estimates of causal effect. For reference, this would resemble confounding scenario (c) (see Methods Figure 5c) with respect confounding structures invalidating assumption GxE2.

Crucially, adopting alcohol and physical activity as interaction covariates yields causal effect estimates which appear biologically plausible, and support evidence from both observational and MR studies suggesting a positive association between BMI and SBP. Estimates using each interaction covariate are presented in Figure 4.

**Figure 4:**
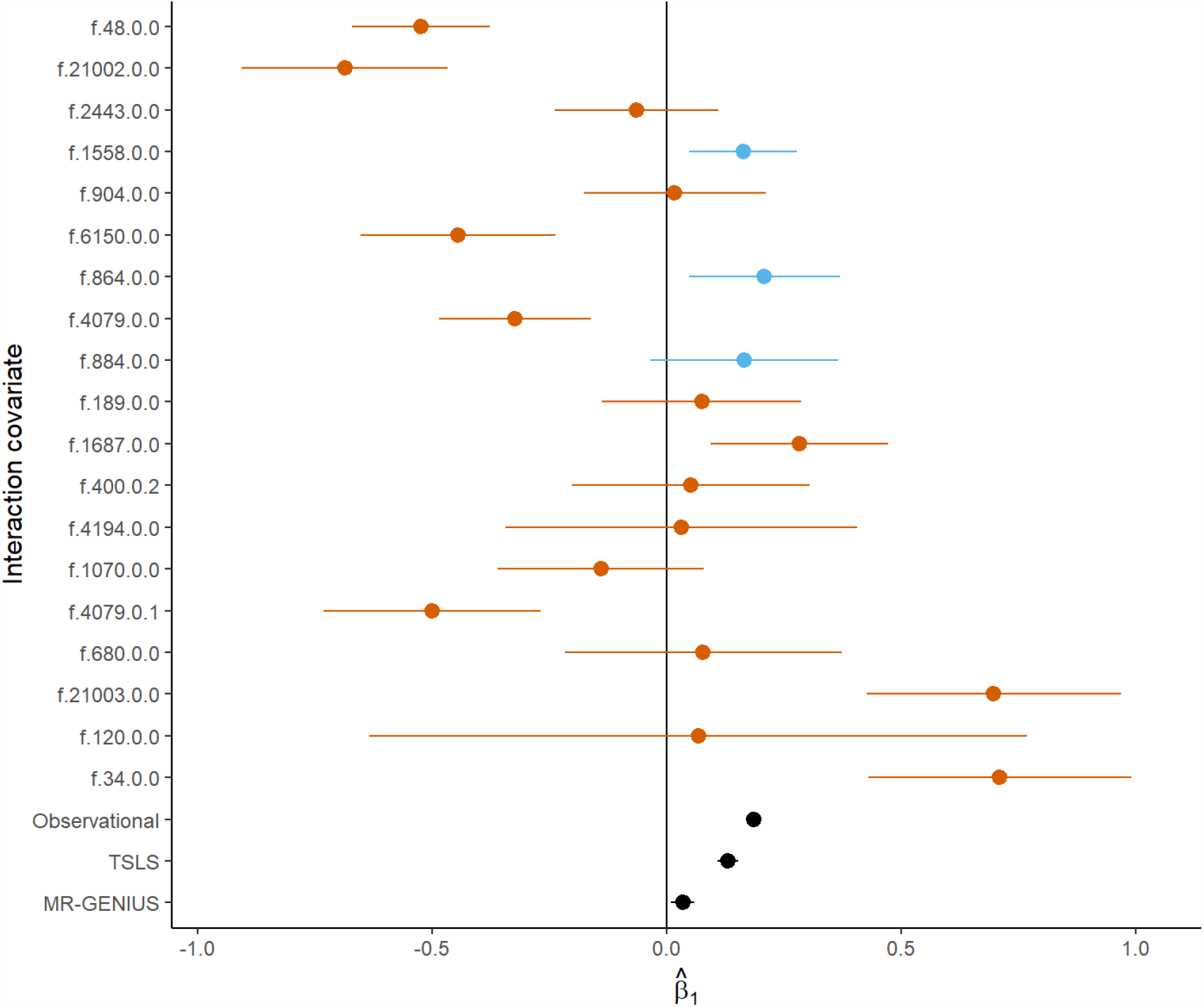
A forest plot showing MR-GxE causal effect estimates using the interaction covariates presented in Table 4. Observation f.4253.0.5 has been omitted for clarity. Red points indicate analyses for which assumptions may likely be violated, while blue points show potentially valid interaction covariates using accompanying sensitivity analyses. Observational, two-stage least squares (TSLS), and MR-GENIUS estimates are also shown as black points.

As a final analysis, we implemented MR-GENIUS using the PRS, BMI, and SBP measures from UK Biobank. This resulted in a more precise estimate in comparison to MR-GxE, however, the effect estimate appears to strongly disagree with evidence from MR-GxE and alternate approaches. Given MR-GENIUS implicitly relies upon analogous assumptions to MR-GxE, it seems reasonable to assume that such a discrepancy could arise from bias due to violations stemming from one or more unmeasured interactions. This is further supported by MR-GxE estimates of similar direction and magnitude which appear to show evidence of bias, such as vigorous physical activity which shows evidence of GxE3 violation.

## Discussion

In this paper we examine two related interaction-based MR approaches: MR-GxE and MR-GENIUS. Both MR-GxE and MR-GENIUS rely upon similar underlying assumptions, whilst differing based on whether a gene-by-covariate interaction needs to be explicitly incorporated within the estimation model. Specifically, MR-GxE relies upon at least a single measured gene-by-covariate interaction which satisfies assumptions GxE 1-3, whilst MR-GENIUS does not require such an interaction to be observed. However, as a consequence of implicitly leveraging multiple underlying interactions, the MR-GENIUS approach requires assumptions GxE 1-3 to hold globally. Essentially, stronger assumptions are required to mitigate the absence of an observed gene-by-covariate interaction.

Through an examination of the MR-GxE assumptions, several approaches aiming to evaluate assumptions GxE 1-3 have been outlined. Interaction strength (GxE1) can be evaluated using the first-stage F-statistic for the interaction term, analogous to evaluating instrument strength in conventional MR. The corresponding global test for interaction strength using MR-GENIUS and a continuous exposure is the Breusch-Pagan test for heteroskedasticity^12, 19^.

Assumption GxE2 can initially be evaluated by estimating the correlation between *Z*_*i*_ and both *G*_*i*_ and *X*_*i*_ respectively. Where *Z*_*i*_ is observed to be correlated with *G*_*i*_, it is possible that a confounding relationship exists violating assumption GxE2. Further, the simultaneous association of *Z*_*i*_ with *G*_*i*_ and *X*_*i*_ can result in bias where *Z*_*i*_ is downstream of *X*_*i*_. However, as the existence of such correlations does not necessarily imply that this assumption is violated, a more promising approach may be to adopt an interaction covariate *Z*_*i*_ which is highly likely to be exogenous (see Methods). For example, one could employ genetic variants which instrument a likely interaction covariate. Future work will explore this possibility.

The constant pleiotropy assumption (GxE3) can be tested in cases where the initial instrument *G*_*i*_ is a composite instrument, that is, comprised of multiple sub-instruments such as genetic variants within a PRS. Heterogeneity in effect estimates obtained using sub-instruments can be considered as evidence of violation of the constant pleiotropy assumption, analogous to heterogeneity in two-sample summary MR^4, 20^. In principle, a similar approach can be applied using sub instruments with MR-GENIUS, though such an examination is beyond the scope of this paper.

In the applied analysis the association between BMI and SBP was estimated using MR-GENIUS and a range of interaction covariates in conjunction with MR-GxE. We identified four suitable interaction covariates, which suggest a positive effect of BMI upon SBP in agreement with previous observational and MR analyses. Importantly, we highlight interaction covariates which violate the MR-GxE assumptions and link these issues to the possibly biased effect estimates obtained using MR-GENIUS.

Several limitations remain with respect to MR-GxE which warrant further explanation. Firstly, reliance upon an observed gene-by-covariate interaction limits the extent to which the method can be applied in contrast to MR-GENIUS. We advocate the use of MR-GENIUS in cases where no interaction covariate is available, though care needs to be taken in justifying the more stringent assumptions MR-GENIUS entails. Second, evaluating GxE2 using the correlation of between *Z*_*i*_ and *G*_*i*_ does not provide a clear indication of whether the assumptions hold. It is possible that GxE2 can be violated when *Z*_*i*_ and *G*_*i*_ appear to be independent, and assuming the direction of effect between *Z*_*i*_ and *X*_*i*_ relies upon a priori knowledge regarding the direction of association. It is therefore critical to identify plausible biological mechanisms underpinning the observed relationships in the MR-GxE model.

Finally, whilst an overidentification test has been presented for evaluating GxE3, there is not at present a method aiming to correct for violation of the constant pleiotropy assumption. It is likely that pleiotropy robust methods, such as median or modal regression, could be utilised to correct for resulting bias, and the application of such methods will be fully explored in future work.

## Methods

### The MR-GxE Approach

The extent to which instrument strength varies across strata of a given interaction covariate *Z*_*k*_ is quantified by the magnitude of the first-stage interaction between *G*_*i*_ and *Z*_*ki*_ (*γ*_3*k*_) shown in equation (3). The MR-GxE approach also relies upon *β*_4*k*_ and *θ*_3*k*_ being equal to zero, analogous to instrument validity in conventional MR. In previous work, MR-GxE was implemented using an approach analogous to MR-Egger regression in two-sample summary MR^4, 7^. This is achieved by initially obtaining strata specific instrument-exposure and instrument-outcome associations, after which the instrument-outcome associations are regressed upon the instrument-exposure associations including an intercept.

The interpretation of estimates and corresponding plots using this approach essentially mirrors MR-Egger regression, though each observation represents a different strata-specific subgroup as opposed to a unique genetic instrument^4^. However, whilst this approach does allow for estimation using publicly available GWAS summary data it has two primary limitations. Initially, it relies upon specification of an interaction-covariate without a readily applicable measure of interaction strength. Second, ambiguity surrounding the definition of interaction-covariate strata can potentially have a substantial impact on causal effect estimates^7^. Applying MR-GxE in an individual level data setting avoids these issues and allows for additional information to be incorporated into effect estimation. Further, such an approach is increasingly applicable with the emergence of large-scale studies with genetic data such as the UK Biobank.

A reduced form model for *Y*_*i*_ given *G*_*i*_ and *Z*_*i*_ incorporating equations (5-6) can be written as

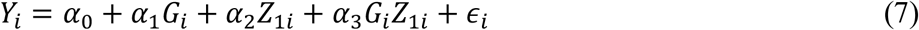

Note that a *G*_*i*_ × *Z*_1*i*_ term is omitted from the second-stage model given in equation (6) due to its role as an instrument, whilst the inclusion of *G*_*i*_ allows for estimation of a horizontal pleiotropic effect on the outcome, denoted by *β*_2_. Using equation (5) and equation (7) we can define the MR-GxE estimand as

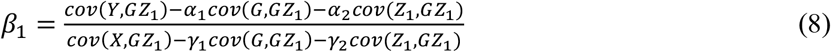

### The MR-GENIUS Approach

The MR-GENIUS approach is an adapted form of Robins’ G-estimation which is robust to additive confounding and pleiotropic bias^12, 21, 22^. This essentially involves leveraging differences in the variance of a given exposure *X*_*i*_ across subgroups of a genetic instrument *G*_*i*_, which are likely the consequence of one or more gene-by-covariate interactions. In the case of a binary instrument and exposure, and using notation from equations 1-5, the MR-GENIUS estimator can be written as:

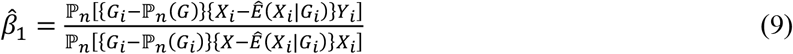

where 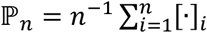 and 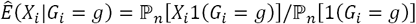^12^.

MR-GENIUS is implemented by first regressing *X*_*i*_ upon *G*_*i*_ and obtaining a set of residuals 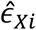. These residuals are then used to create an instrument 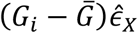 which is incorporated within a TSLS model, as a single instrument for *X*_*i*_^12^. Estimates of 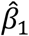 remain unbiased, provided the instrument *G*_*i*_ is associated with the exposure of interest, the effect does not change across values of the unmeasured confounders, and the MR-GENIUS model is identified such that the change in variance across levels of the instrument is non-zero^12^. In the binary exposure case, this means that the MR-GENIUS model is identified when *cov*(*G*_*i*_, *var*(*X*_*i*_|*G*_*i*_)) ≠ 0, and for a continuous exposure the MR-GENIUS model when the residual error *ϵ*_*X*_ is heteroskedastic, that is, not constant across levels of *G*_*i*_^12^. This can be evaluated using a Breusch-Pagan test for heteroskedasticity^12, 19^.

These conditions also restrict the degree of joint effect modifiers of both *X*_*i*_ and *Y*_*i*_. Importantly, it should be noted that the interaction covariate need not be explicitly identified using MR-GENIUS, illustrated by the absence of any *Z*_*ki*_ in equation 9. However, identification of the MR-GENIUS model implicitly relies upon the presence of one or more gene-by-covariate interactions to induce the desired dependence between *G*_*i*_ and *var*(*X*|*G*).

### GxE1: Interaction strength

The MR-GxE estimator can be viewed as an extension of the Wald ratio, including an adjustment for the direct effects of *G*_*i*_ and *Z*_*ki*_. Thus, in the special case where *G*_*i*_ and *Z*_*ki*_ are marginally independent of the exposure and outcome (but their interaction via a single covariate *Z*_*ki*_ is not), the MR-GxE estimator collapses to:

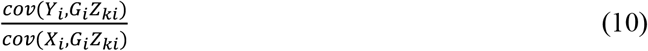

MR-GxE is clearly reliant upon a strong first-stage interaction, such that *γ*_*k*3_ ≠ 0 in order to make the denominator of (10) non-zero (GxE1). As in the conventional MR setting it is possible to assess interaction strength using the F-statistic for the interaction term in the first-stage model, however, the use of a single instrument precludes relating the F-statistic to the magnitude of relative bias (at least three instruments would be required in this case for the asymptotic formula to be valid). Therefore, whilst an F-statistic of 10 may satisfy the standard threshold for sufficient instrument strength, it is not possible to relate this to a 10% relative bias towards the OLS estimate. Further, as in the conventional MR setting, interaction strength does not mitigate bias from violations of assumptions GxE 2-3^23^. Where possible, candidate interactions should be identified in separate samples to avoid issues related to Winner’s curse, as is the case with instrument selection in conventional MR.

At this point it is important to highlight several features of gene-by-covariate interactions which require careful consideration prior to performing MR-GxE. Firstly, interactions are scale dependent, and as a result applying transformations can create spurious associations^24^. As such spurious associations can exist as an artefact of the data, estimates leveraging such information can potentially be unreliable. A related concern is that the interaction may not be linear, as is assumed in equation 5. A potential solution to this issue is to fit flexible models (e.g., fractional polynomial models, which include varying exponents with respect to *G*_*i*_*Z*_*ki*_) to allow for non-linear interactions to be identified^25^.

As MR-GENIUS does not require gene-by-covariate interactions to be identified, testing for identification is performed globally by evaluating heteroskedasticity with respect to the residuals *ϵ*_*Xi*_. Specifically, MR-GENIUS relies upon the residual error in a regression of the exposure upon the instrument to be heteroskedastic, such that 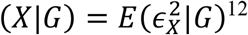.

### GxE2: Interaction exogeneity

In previous work we show how assumption GxE2 is most likely violated when certain confounding structures exist, specifically, where *G*_*i*_ and *Z*_*i*_ are simultaneously downstream of a confounder *U*_*i*_ or where there is an open path between the two variables through *U*_*i*_^7^. These confounding structures are illustrated in Figure 5, where scenarios (a), (b), and (d) represent pathways which can result in bias. In contrast, scenario (c) represents a blocked path which does not necessarily result in bias.

**Figure 5:**
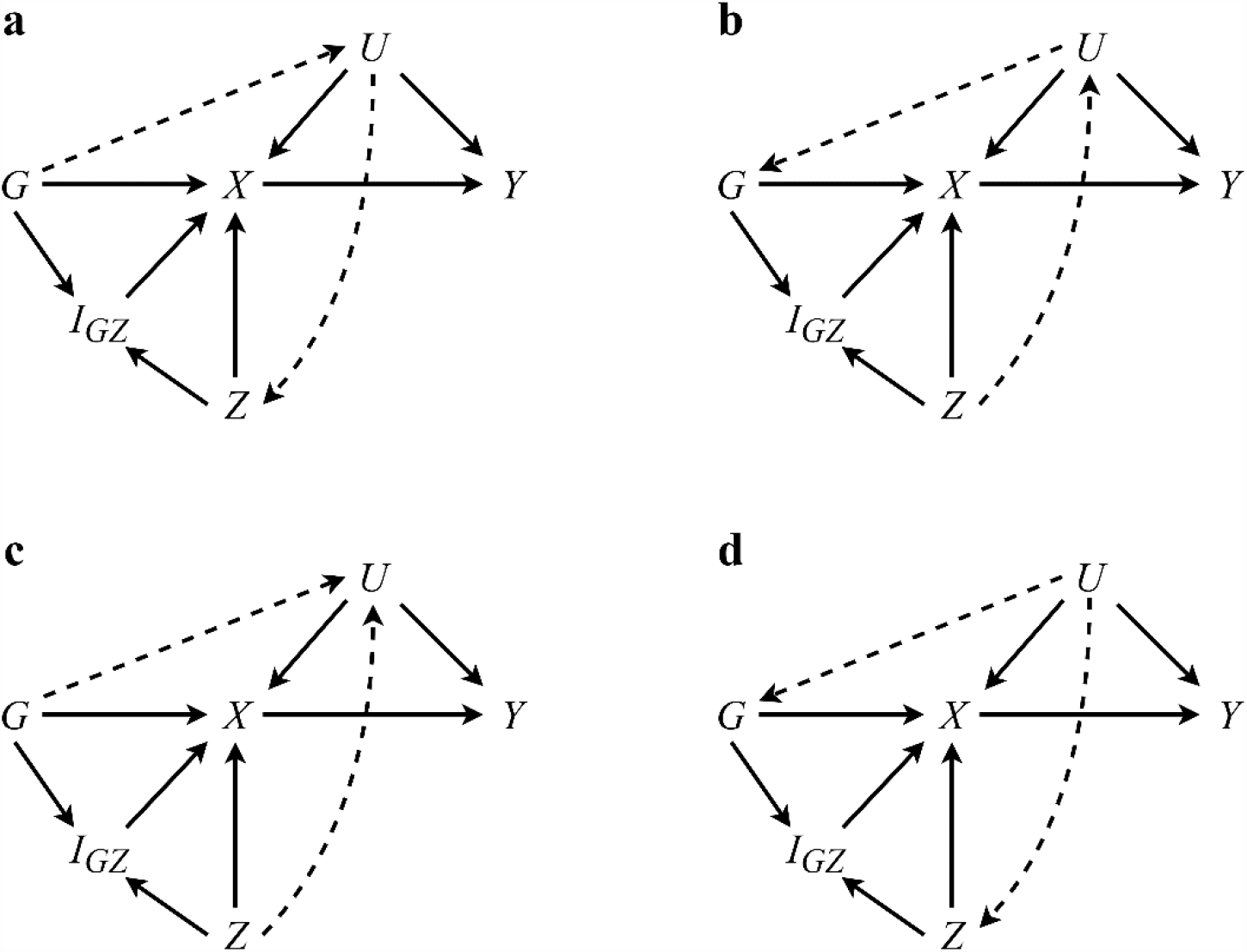
A panel showing a set of 4 DAGs wherein the instrument *G* and interaction covariate *Z* are simultaneously associated with one or more confounders *U*. In this case, DAGs (a), (b), and (d) would likely result in biased causal effect estimates using MR-GxE, in contrast to confounding structure (c).

To understand why scenarios (a)-(c) induce bias into MR-GxE estimates, we can extend the MR-GxE estimand to allow for violation of GxE 2, by including covariance terms between *U*_*i*_ and (*Z*_*i*_, *G*_*i*_*Z*_*i*_)*U*_*i*_, such that were it possible to include *U*_*i*_ in the TSLS model, the resulting estimate could be written as:

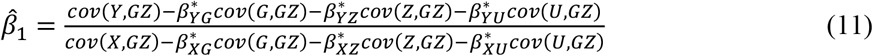

where each *β*^∗^indicates a multivariable regression estimate pertaining to the second subscript variable when regressed upon the first, including the unmeasured confounder *U*_*i*_. As it is not possible to directly measure and adjust for *U*_*i*_, we rely upon independence between *U*_*i*_ and *G*_*i*_*Z*_*i*_ for equation (11) to be equivalent to the MR-GxE estimator in equation (8).

Violation of GxE2 and subsequent bias can result from specific configurations of *G*_*i*_, *Z*_*i*_, and *U*_*i*_ associations. For example, a pathway from *G*_*i*_ to *Z*_*i*_ through *U*_*i*_ (see Figure 5a) would result in bias, though the converse is less likely where genetic variants are used as instruments^7^.

A more serious concern is the potential for collider bias when estimating fitted values 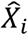 in the first-stage MR-GxE model. As shown in equation (5), it is necessary to include the interaction covariate in the first-stage model. However, in cases where *G*_*i*_ and *U*_*i*_ are both simultaneously upstream associated with *Z*_*i*_, conditioning on *Z*_*i*_ will induce collider bias in the first-stage MR-GxE model, such that the estimate of pleiotropic effect 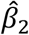 and subsequent adjustment will be inaccurate. This case is illustrated in Figure 6.

**Figure 6:**
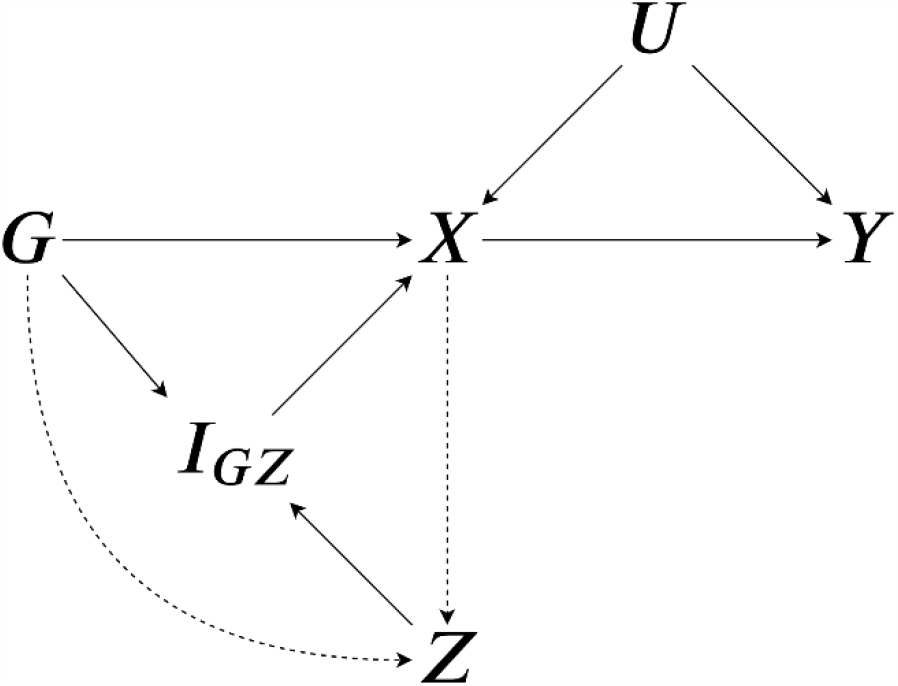
A DAG showing a situation in which conditioning on *Z* when *G* and *X* are simultaneously upstream associated with *Z* would induce collider bias in a regression of *X* upon *G* and *Z* outlined in equation 5.

We present two strategies for limiting the impact of such bias. As an initial test, it is possible to estimate the correlation between *G*_*i*_ and *Z*_*i*_, with observed independence serving as evidence against violation of GxE2. Specifically, with reference to Figure 5 scenarios (a), (b), and (d) can in principle lead to observed correlations between *G*_*i*_ and *Z*_*i*_ which can be identified. However, it is important to emphasise that independence cannot necessarily be interpreted as GxE2 being satisfied. This would primarily be the case where a three-way interaction exists between the instrument *G*_*i*_, interaction covariate *Z*_*i*_, and one or more confounders *U*_*i*_. Rather than removing the possibility, an observed correlation between *G*_*i*_ and *Zi* can highlight a potential issue in the analysis which warrants further consideration.

A second and potentially more robust approach would be to adopt a genetic proxy variable for the interaction covariate *Z*_*i*_, as this would share the same benefits with regard to causal direction as *G*_*i*_ with respect to environmental confounders. For example, when estimating the association between alcohol and SBP using education as an interaction covariate, adopting a PRS for education would in principle utilise the explained variation in education excluding environmental confounders such as socio-economic status.

### GxE3: Constant pleiotropy

The third MR-GxE assumption requires pleiotropic effects of *G*_*i*_ upon *Y*_*i*_ to remain constant across values of *Z*_*i*_, with the gene-by-covariate interaction being independent of *Y*_*i*_ when conditioning on *X*_*i*_. Where this is not the case estimates of causal effect will exhibit bias, such that the degree of bias in the MR-GxE estimate will be equal to:

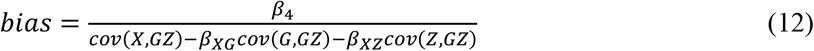

To illustrate how over-identification tests can be applied in the context of MR-GxE, consider an extension of equations 5 and 6 to include an arbitrary number of sub-instruments, wherein a single instrument ***G***_***i***_ is comprised of *m* ∈ {1,2, …, *M*} sub-instruments. Where *G*_*mi*_ denotes the *m*^*th*^ sub-instrument in ***G***_***i***_, we can rewrite the data generating models presented in equations (3-4) as:

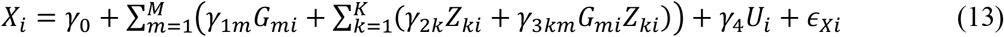

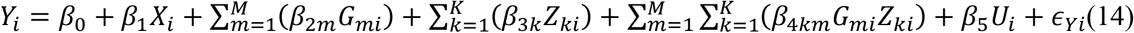

Sargan tests can then be used to compare different MR-GxE estimates of the same causal parameter provided we have more instruments of sufficient strength than we need to consistently estimate the parameter^14^.

## Data Availability

UK Biobank data was used, accessed through application number 8786. For more information, please visit https://www.ukbiobank.ac.uk/

## Acknowledgements

The authors would like to thank Professor Frank Windmeijer and Dr Zoltán Kutalik for providing valuable comments and suggestions. UK Biobank analyses were conducted using application number 8786.

